# CSF sphingomyelins in Alzheimer’s disease, neurodegeneration, and neuroinflammation

**DOI:** 10.1101/2022.01.19.22268636

**Authors:** Autumn Morrow, Daniel J. Panyard, Yuetiva K. Deming, Erin Jonaitis, Ruocheng Dong, Eva Vasiljevic, Tobey J Betthauser, Gwendlyn Kollmorgen, Ivonne Suridjan, Anna Bayfield, Carol A. Van Hulle, Henrik Zetterberg, Kaj Blennow, Cynthia M. Carlsson, Sanjay Asthana, Sterling C. Johnson, Corinne D. Engelman

**Affiliations:** Department of Population Health Sciences, University of Wisconsin-Madison, 610 Walnut Street, 707 WARF Building, Madison, WI 53726, United States of America; Department of Genetics, School of Medicine, Stanford University, 291 Campus Drive, Stanford, CA 94305, United States of America; Wisconsin Alzheimer’s Disease Research Center, University of Wisconsin-Madison, 600 Highland Avenue, J5/1 Mezzanine, Madison, WI 53792, United States of America; Department of Medicine, School of Medicine and Public Health, University of Wisconsin-Madison, 1685 Highland Avenue, 5158 Medical Foundation Centennial Building, Madison, WI 53705, United States of America; Wisconsin Alzheimer’s Institute, UW School of Medicine and Public Health, 610 Walnut Street, 9th Floor, Madison, WI 53726; Center for Demography of Health and Aging, University of Wisconsin-Madison, 1180 Observatory Drive, Madison, WI 53706; Roche Diagnostics GmbH, Nonnenwald 2, 82377 Penzberg, Germany; Roche Diagnostics International Ltd, Forrenstrasse 2, 6343 Rotkreuz, Switzerland; Institute of Neuroscience and Physiology, The Sahlgrenska Academy at University of Gothenburg, 43180 Mölndal, Sweden; Clinical Neurochemistry Laboratory, Sahlgrenska University Hospital, 43180 Mölndal, Sweden; United Kingdom Dementia Research Institute at UCL, London, WC1E6BT, United Kingdom; Department of Neurodegenerative Disease, UCL Institute of Neurology, London, WC1H0AL, United Kingdom; Hong Kong Center for Neurodegenerative Diseases, Hong Kong, China; William S. Middleton Memorial Veterans Hospital, 2500 Overlook Terrace, Madison, WI 53705, United States of America

**Keywords:** Metabolomics, Alzheimer’s disease, biomarkers, sphingomyelin, sphingolipid, neurodegeneration, neuroinflammation, cerebrospinal fluid

## Abstract

**INTRODUCTION:** Sphingomyelin (SM) levels have been associated with Alzheimer’s disease (AD), but the association direction has been inconsistent and research on cerebrospinal fluid (CSF) SMs has been limited by sample size, breadth of SMs examined, and diversity of biomarkers available.

**METHODS:** Leveraging two longitudinal AD cohorts with metabolome-wide CSF metabolomics data (n=502), we analyzed the relationship between the levels of 12 CSF SMs, and AD diagnosis and biomarkers of pathology, neurodegeneration, and neuroinflammation using logistic, linear, and linear mixed effects models.

**RESULTS:** No SMs were significantly associated with AD diagnosis, mild cognitive impairment, or amyloid biomarkers. Phosphorylated tau, neurofilament light, α-synuclein, neurogranin, soluble triggering receptor expressed on myeloid cells 2, and chitinase-3-like-protein 1 were each significantly, positively associated with at least 5 of the SMs.

**DISCUSSION:** The associations between SMs and biomarkers of neurodegeneration and neuroinflammation, but not biomarkers of amyloid or diagnosis of AD, point to SMs as potential biomarkers for neurodegeneration and neuroinflammation that may not be AD-specific.

## 1. Background

To discover new therapeutic targets for Alzheimer’s disease (AD), we must better understand the changes caused by the disease. One approach to better understanding these changes in AD is metabolomics, a way of studying the byproducts of the body’s metabolic processes. Metabolomics grants a window into the metabolic state of the body[1] and has proven useful in studying a variety of diseases, including identifying a mechanism of insulin resistance for type 2 diabetes, developing precision medicine approaches to treating cancer, and identifying altered metabolites and their mechanism of change in central nervous system-related disorders such as simian immunodeficiency virus[2–5]. Metabolomics has also been a valuable tool for studying AD, as it has helped to identify new biomarkers and mechanisms for the disease[6].

One of the findings from AD metabolomics studies has been the association between the sphingolipid metabolic pathway and AD[7]. Sphingolipids are a family of membrane lipids that participate in diverse and fundamental cellular processes, such as cell division, differentiation, and death[8]. In mammals, sphingomyelins (SMs) are the most abundant molecule of the sphingolipid metabolic pathway (Supplemental Figure 1)[8]. Despite the identification of SMs as associated with AD, there is disagreement as to how they are associated with the disease: some studies show that SMs decrease in tissue across brain regions and in plasma during progression to AD, while others suggest that they increase[9,10]. Previous research in cerebrospinal fluid (CSF) SMs has been limited by sample size, breadth of SMs examined, and diversity of biomarkers available for AD, neurodegeneration, and neuroinflammation, which may have contributed to the lack of clarity in the role of SMs.

Here, we build on our understanding of the role of SM metabolites in AD. Leveraging two large, longitudinal cohorts with metabolome-wide CSF metabolomics, robust cognitive diagnoses, and a diversity of CSF biomarker and brain imaging measures, we analyzed the relationship between SMs, AD diagnosis, and biomarkers of pathology, and markers of neurodegeneration and neuroinflammation. The results shed light on the role of SMs in neurodegeneration and neuroinflammation.

## 2. Methods

### 2.1. Study Cohorts

Data were included from the Wisconsin Registry for Alzheimer’s Prevention (WRAP) and the Wisconsin Alzheimer’s Disease Research Center (ADRC) cohorts[11,12]. These longitudinal studies of preclinical and clinical AD in middle to older aged adults include CSF metabolomics, clinical diagnosis, neuroimaging, and CSF biomarkers of AD, neurodegeneration, and neuroinflammation. All participants included in the current research had at least one lumbar puncture (LP); the CSF samples for WRAP and the Wisconsin ADRC were collected and analyzed following the same protocols. A committee of dementia specialists determined diagnosis of dementia-AD (n = 44 individuals), mild cognitive impairment (MCI) (n = 40 individuals), or cognitively unimpaired (CU) (n = 409 individuals) (Table 1). This study was approved by the University of Wisconsin Health Sciences Institutional Review Board as part of the Generations of WRAP (GROW) study. Participants in the WADRC and WRAP studies provided written informed consent.

**Table 1.** Baseline characteristics of individuals.

| Characteristic | Diagnosis data set<br>(cross-sectional)<br>(n = 493 individuals) | Biomarker data set individuals<br>(n = 494 individuals) | Biomarker data set visits<br>(longitudinal)<br>(n = 726 visits) | Overlap<br>(n = 485 individuals) |
| --- | --- | --- | --- | --- |
| Diagnosis, n (%) |  |  |  |  |
| Dementia-AD | 44 (8.9%) | 43 (8.7%) | 44 (6.1%) | 43 (8.9%) |
| MCI | 40 (8.1%) | 42 (8.5%) | 45 (6.2%) | 40 (8.2%) |
| CU | 409 (82.9%) | 409 (82.8%) | 637 (87.7%) | 402 (82.9%) |
| Primary Race, n (%) |  |  |  |  |
| White | 472 (95.7%) | 473 (95.7%) | 693 (95.5%) | 464 (95.7%) |
| Black or African American | 16 (3.3%) | 16 (3.2%) | 22 (3.03%) | 16 (3.3%) |
| American Indian or Alaska Native | 3 (0.6%) | 3 (0.6%) | 4 (0.6%) | 3 (0.6%) |
| Asian | 1 (0.2%) | 1 (0.2%) | 3 (0.4%) | 1 (0.2%) |
| Other | 1 (0.2%) | 1 (0.2%) | 4 (0.6%) | 1 (0.2%) |
| Sex, n (%) |  |  |  |  |
| Male | 191 (38.7%) | 191 (38.7%) | 273 (39.4%) | 189 (39.0%) |
| Female | 302 (61.3%) | 303 (61.3%) | 453 (65.4%) | 296 (61.0%) |
| CSF amyloid and tau status, n (%) |  |  |  |  |
| A-T- | 319 (64.7%) | 324 (65.6%) | 500 (68.9%) | 317 (65.4%) |
| A+T- | 61 (12.4%) | 59 (11.9%) | 85 (11.7%) | 59 (12.2%) |
| A-T+ | 20 (4.1%) | 20 (4.0%) | 28 (3.8%) | 19 (3.9%) |
| A+T+ | 85 (17.2%) | 84 (17.0%) | 102 (14.0%) | 83 (17.1%) |
| Not measured | 8 (1.6%) | 7 (1.4%) | 11 (1.5%) | 7 (1.4%) |
| Age in years mean, (SD) | 64.1 (8.9) | 63.1 (8.9) | 63.4 (8.3) | 64.0 (9.0) |
| PiB Global DVR, n, (SD) | 179 (9.4) | N/A | N/A | 179 (9.4) |
| PiB Positive, n, (%) | 50 (27.9%) | N/A | N/A | 50 (27.9%) |

Abbreviations: AD, Alzheimer's disease; MCI, mild cognitive impairment; CU, cognitively unimpaired; CSF, cerebrospinal fluid; PiB, Pittsburgh Compound B; DVR, distribution volume ratio; SD, standard deviation.

### 2.2. CSF samples, biomarkers, and metabolomics

The process through which CSF samples were acquired and biomarker concentrations were measured has been previously described[13]. Briefly, CSF samples were collected in the morning after fasting. Within 30 minutes of collection, samples were mixed, centrifuged, aliquoted, and then stored at −80°C. All CSF sample biomarker assays were performed at the Clinical Neurochemistry Laboratory, University of Gothenburg from March 2019 to January 2020. All biomarker data were taken from the Roche NeuroToolKit (NTK; Roche Diagnostics International Ltd, Rotkreuz, Switzerland), a panel of exploratory prototype assays designed to robustly evaluate biomarkers associated with key pathologic events characteristic of AD and other neurological disorders, as previously described[13]. For metabolomic analyses, CSF samples were shipped overnight to Metabolon, Inc. (Durham, NC), where samples were also kept frozen at −80°C until analysis[14]. The untargeted metabolomics analysis was performed using Ultrahigh Performance Liquid Chromatography-Tandem Mass Spectrometry (UPLC-MS/MS). Chemical properties, metabolite identifiers, and pathway information were provided for each metabolite. All metabolite data underwent a quality control (QC) process. Of the 412 metabolites in the initial sample, 13 were removed for being missing in ≥ 50% of the samples. Nine metabolites were removed for low variance (interquartile range = 0). Of the 1,172 CSF samples, one was removed for missing ≥ 40% of the metabolite values. We also removed 220 samples obtained as part of a clinical trial (CT: 00939822). Metabolite values were log_10_ transformed to avoid skewness. After these QC steps, data were available on 390 metabolites from 951 CSF samples taken from 502 individuals. Of the 390 metabolites, 12 metabolites in the sphingolipid metabolic pathway were used in the analyses described. Of the 12 metabolites included in these analyses, 9 were designated by metabolon as tier 2 compounds, meaning that these compounds have a structure that has been confirmed by literature review, but they are not necessarily confirmed by a reference standard. These 9 metabolites were: behenoyl SM (d18:1/22:0), palmitoyl dihydro-SM (d18:0/16:0), SM (d18:1/14:0, d16:1/16:0), SM (d18:1/20:0, d16:1/22:0), SM (d18:1/20:1, d18:2/20:0), SM (d18:1/22:1, d18:2/22:0, d16:1/24:1), SM (d18:1/24:1, d18:2/24:0), SM (d18:2/16:0, d18:1/16:1), SM (d18:2/24:1, d18:1/24:2).

### 2.3. Neuroimaging

Detailed methods for radiotracer synthesis and positron emission tomography (PET) and magnetic resonance imaging (MRI) data acquisition, processing, and quantification have been previously described[15]. Briefly, anatomical MRI (T1-w and T2-w) underwent multispectral unified tissue class segmentation (SPM12)[16]. Regions of interest (ROIs) for PET analysis were defined by applying the inverse deformation field defined during tissue segmentation to the MNI152-space Automated Anatomical Labeling atlas [17] and restricting the subject-space ROIs to voxels with gray matter probabilities greater than 0.3. Reconstructed dynamic Pittsburgh compound B (PiB) PET data acquired from 0-70 minutes post nominal 555 MBq [^11^C] PiB injection on a Siemens EXACT HR+ or Siemens Biograph Horizon PET/CT were isotopically smoothed, interframe realigned, dynamically denoised, and registered to T1-weighted MRI[15]. Amyloid burden was assessed by averaging distribution volume ratio (DVR) estimates across eight bilateral regions (Logan graphical analysis, cerebellum gray matter reference region, k2’=0.149 min^-1^; ROIs included angular gyrus, anterior and posterior cingulate, medial orbital-frontal gyrus, precuneus, supramarginal gyrus, and middle and superior temporal gyri)[18]. The resulting measurement is referred to as the PiB Global DVR.

### 2.4. Data Integration

CSF data were matched to diagnosis data from the nearest clinic visit. To remove potential correlation between genetically related participants, only the oldest individual from each family group was selected (n = 32 individuals removed). Data were divided into two overlapping data sets for analysis. First was the cross-sectional “Diagnosis” data set, which was focused on clinical diagnoses, comprising the oldest visit per participant (n = 493), diagnosis at that visit, and matched CSF metabolomics and PiB data for that visit. Logistic regressions were used to identify associations between diagnosis and SMs, and linear regressions were used for associations between PiB Global DVR and SMs. SM-PiB Global DVR analyses were cross-sectional because there were not enough PiB Global DVR measurements per individual to perform a longitudinal analysis. However, results for these analyses with PiB as the outcome were presented with the results from the AD biomarkers (data set described below) to better group the outcomes conceptually.

The second data set was the longitudinal “Biomarker” data set, which focused on CSF biomarkers, comprising all available visits with CSF biomarker data and their corresponding metabolomics data (n = 494 individuals and n = 726 samples). This data set included all available LP visits for each individual to maximize sample size (Table 1). The Biomarker data set was used to identify associations between CSF biomarkers and metabolites. AD biomarkers included CSF Aβ_42_/Aβ_40_, CSF p-tau_181_/Aβ_42_, and CSF p-tau_181_[19–21]. The neurodegeneration biomarkers used were neurogranin, neurofilament light (NfL), and alpha-synuclein (α-synuclein)[22–24]. We also included biomarkers for neuroinflammation: interleukin-6 (IL6), chitinase-3-like protein 1 (YKL40), and soluble triggering receptor found on myeloid cells 2 (sTREM2)[25–27]. All Biomarker data were checked for skewness, and NfL, p-tau_181_, p-tau_181_/Aβ_42_, and IL6 were log_10_-transformed for skewness ≥ 2[28].

### 2.5. Data Analysis

All analyses were performed using R (version 4.0.2) and the “Tidyverse” packages (version 1.3.0)[29]. Logistic and linear regressions were performed using the glm and lm functions, respectively, from the “stats” package (version 3.6.2)[30]. Linear mixed effects models (LMMs) using the biomarker data set were performed using the lmer function from the “lme4” and “lmerTest” packages[31,32]. All P-values from regression models were subjected to a Bonferroni adjusted threshold of P ≤ 3.47×10^−4^ to determine significance (α = 0.05 / 12 metabolites / 12 primary outcomes).

#### 2.5.1. SM association with dementia-AD and MCI diagnoses

In the Diagnosis data set, diagnoses of dementia-AD and MCI (both relative to cognitively unimpaired [CU]) were separately regressed on each metabolite, controlling for sex and age. Pseudo R^2^ values to evaluate model fit were calculated for each regression using the R2 function from the “semEff” package (version 0.4.0)[33].

#### 2.5.2. SM association with biomarkers of AD, neurodegeneration, and neuroinflammation

For PiB Global DVR, cross-sectional associations with SMs were analyzed using the Diagnosis data set and a linear regression model adjusting for sex and age. Adjusted R^2^ values were calculated for the model, to assess model fit.

LMMs were used to determine the association between SMs and biomarkers for which longitudinal data were available (Aβ_42/40_, p-tau_181_/Aβ_42_, p-tau_42_, neurogranin, NfL, α-synuclein, IL6, YKL40, and sTREM2) in the Biomarker data set. Models were adjusted for age, sex, and a random intercept for the participant ID to account for multiple observations per individual. Marginal R^2^ values for these models were calculated using the r.squaredGLMM function from the “MuMIn” package (version 1.43.17) to assess model fit[34].

#### 2.5.3. Independent signals

To assess the extent to which SMs represented the same underlying signal, pairwise correlations were calculated between all 72 pairs of SMs. To determine whether models including the most strongly associated SM from our main analyses as a predictor were significantly improved by adding any other SMs, we repeated the main analyses with the top SM (stearoyl SM) as the main predictor, adding each of the other metabolites as an additional predictor.

We used an analysis of variance (ANOVA) test to assess whether the model with stearoyl SM was significantly different from the model that included both stearoyl SM and an additional SM. We used the anova function from the “stats” package (version 4.0.2) to compare each pair of nested models[30]. To ensure equal sample sizes between the two groups, samples missing any of the values necessary for either regression were dropped. We compared the R^2^ values calculated for each of the regressions before and after removing the necessary samples to perform this analysis to ensure that they were not drastically altered by these removed samples.

#### 2.5.4. Sensitivity Analyses

*APOE* ε2/ε3/ε4 genotype was determined using competitive allele-specific PCR-based KASP genotyping for rs429358 and rs7412[14]. Because *APOE* ε4 count is strongly associated with AD and thus may feasibly influence our results, we performed a sensitivity analysis in which we repeated the main analyses but also controlled for the number of C alleles (0, 1, or 2) at the rs429358 SNP, which effectively quantifies the number of ε4 alleles[35]. Only participants whose self-reported race was “white” were retained for this analysis because of previously reported heterogeneity in effect of the *APOE* ε4 allele by race[36]. We added *APOE* ε4 count as a covariate for these analyses.

Some studies of SMs in plasma have suggested that the direction of association of SMs and AD differ based on sex[37–39]. To determine whether there are sex-specific differences in associations with SMs in CSF, the SM-diagnosis and SM-biomarker regression analyses were repeated with interaction effects modeling, where the original regression models were rerun with the addition of a term for SM*female. A second sensitivity analysis was also conducted with stratification by sex (see Supplemental Table 1 for sex-stratified sample characteristics). Similarly, to understand whether associations between SMs and biomarkers of AD, neurodegeneration, and neuroinflammation are different with amyloid and tau positivity, we performed the SM-diagnosis and SM-biomarker regressions with additional terms for A+T+ (binary indicator), A+T-(binary indicator), SM*A+T+, and SM*A+T-. Using thresholds that were previously defined in the WRAP study, amyloid-positive individuals were defined as those with Aβ_42/40_ values below 0.046 pg/mL, and tau-positive individuals were defined as those with p-tau_181_ values above 24.8 pg/mL[13]. We additionally then conducted an analysis stratified by amyloid and tau status, in which the main SM-biomarker regressions were performed for all individuals who were amyloid-positive and tau-positive (A+T+), amyloid-positive and tau-negative (A+T-), and amyloid-negative and tau-negative (A-T-) separately (see Supplemental Table 2 for amyloid and tau status-stratified sample characteristics)[40].

Each SM-diagnosis and SM-biomarker regression was repeated as in the main set of regressions with the noted change for each sensitivity analysis. Regression results were subjected to the same Bonferroni-corrected significance threshold used in the main analyses (P < 3.47×10^−4^). P-values and effect sizes from the sensitivity analyses were compared to the results of the main set of analyses.

## 3. Results

In both data sets, most participants were CU, white, female, and amyloid- and tau-negative as measured by LP (Table 1). The average baseline ages in the Diagnosis and Biomarker data sets were 64.1 (SD = 8.9) and 63.1 (SD = 8.9) years, respectively. Metabolite and biomarker missingness is described in Supplemental Table 3.

### 3.1. Pairwise correlations

Pairwise correlations between metabolites showed that the metabolites were all closely correlated (Figure 1). No negative correlations were observed; correlation coefficients ranged from 0.46 to 0.89 (mean = 0.74); behenoyl SM (d18:1/22:0) and stearoyl SM (d18:1/18:0) were the least correlated of all metabolite pairs (Supplemental Table 4).

**Figure 1.**
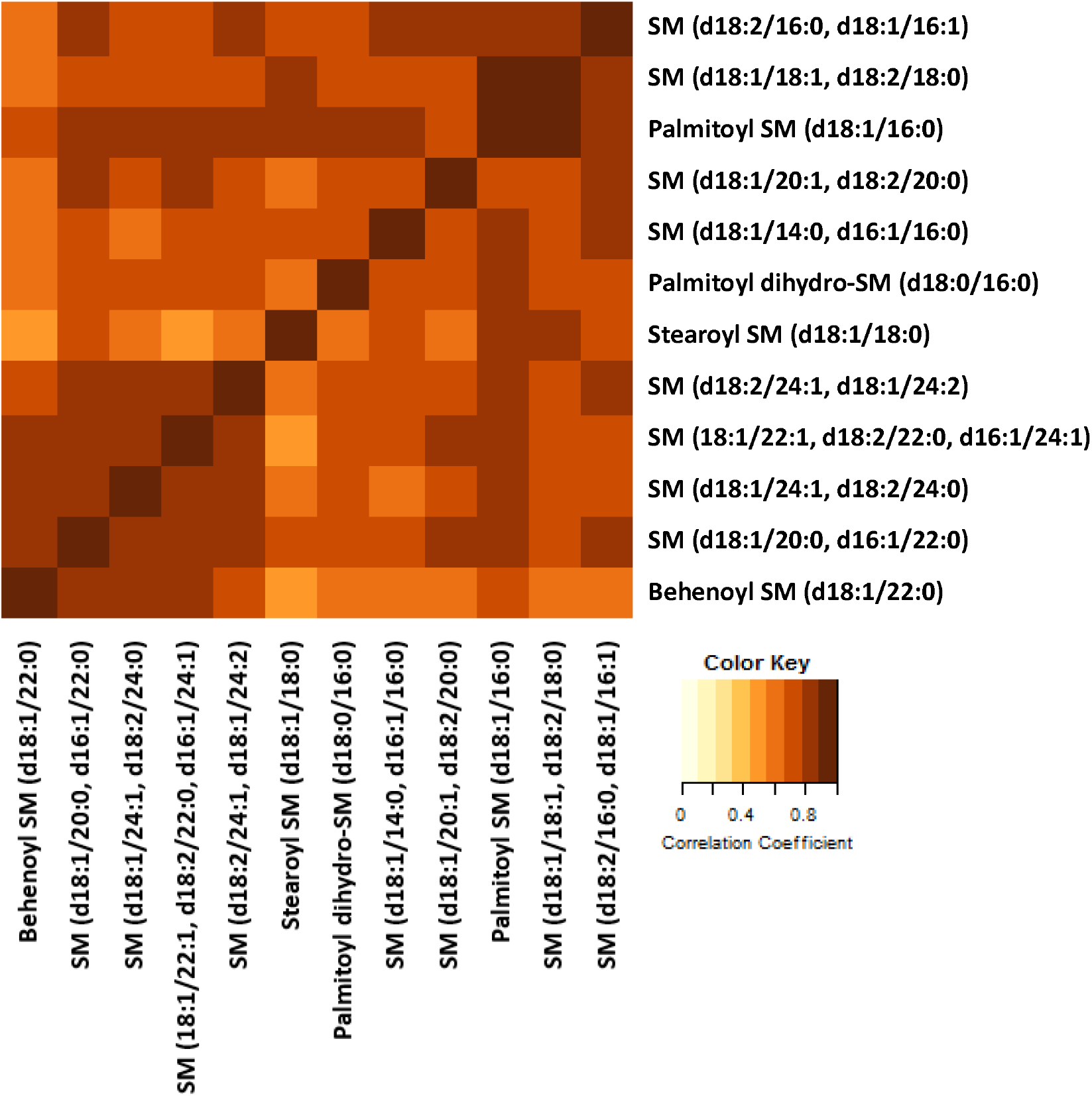
Pairwise correlations of SMs. A heatplot showing the correlation between each metabolite pair. Darker colors indicate stronger correlations, while lighter colors indicate weaker correlations. The lowest correlation coefficient was 0.47 and the highest was 0.89.

### 3.2. SM associations with dementia-AD

Of the SMs analyzed, none showed a significant association with either dementia-AD or MCI diagnosis relative to CU controls (Supplemental Table 5).

### 3.3. SM associations with biomarkers of AD, neurodegeneration, and neuroinflammation

SM associations with AD biomarkers (PiB Global DVR, CSF biomarkers of Aβ_42/40_, p-tau_181_/Aβ_42_, and p-tau_181_) were substantially different between measures of amyloid and tau. None of the 12 metabolites were significantly associated with any amyloid-related biomarkers (Aβ_42/40_, PiB Global DVR, and p-tau_181_/Aβ_42_) after Bonferroni correction (Table 2). Scatterplots for these outcomes plotted against metabolite levels showed large clusters of data points with no clear pattern (Figure 2B). In contrast, 12 SMs had positive, nominally significant associations with CSF p-tau_181_ (P < 0.05), and 6 associations remained significant after Bonferroni correction. Scatterplots of stearoyl SM (d18:1/18:0), the most significantly associated metabolite, and p-tau_181_ reflected this positive association (Figure 2B).

**Table 2.** Associations between SMs and AD biomarkers.

| | PiB Global DVR | | | A$\beta_{42/40}$ | | | p-tau<sub>181</sub>/A$\beta_{42}$ (pg/mL) | | | p-tau<sub>181</sub> (pg/mL) | | |
| --- | --- | --- | --- | --- | --- | --- | --- | --- | --- | --- | --- | --- |
| Metabolite | $\beta$ | P | R<sup>2</sup> | $\beta$ | P | R<sup>2</sup> | $\beta$ | P | R<sup>2</sup> | $\beta$ | P | R<sup>2</sup> |
| behenoyl SM (d18:1/22:0) | 0.02 | 0.85 | 0.09 | -3.54E-04 | 0.86 | 0.13 | 0.02 | 0.54 | 0.16 | 0.02 | 0.07 | 0.13 |
| palmitoyl dihydro-SM<br>(d18:0/16:0) | 0.07 | 0.60 | 0.09 | -3.00E-04 | 0.93 | 0.13 | 0.01 | 0.84 | 0.15 | 0.04 | 0.04 | 0.14 |
| palmitoyl SM (d18:1/16:0) | -0.19 | 0.23 | 0.10 | -1.87E-03 | 0.68 | 0.14 | 0.03 | 0.64 | 0.16 | **0.17** | **5.59E-10** | **0.16** |
| SM (d18:1/14:0, d16:1/16:0) | -0.05 | 0.70 | 0.09 | -1.41E-03 | 0.68 | 0.14 | 0.03 | 0.58 | 0.16 | **0.09** | **8.09E-07** | **0.14** |
| SM (d18:1/18:1, d18:2/18:0) | -0.15 | 0.38 | 0.10 | -3.22E-03 | 0.50 | 0.14 | 0.07 | 0.30 | 0.16 | **0.22** | **5.10E-13** | **0.17** |
| SM (d18:1/20:0, d16:1/22:0) | 0.01 | 0.93 | 0.09 | -2.60E-03 | 0.38 | 0.14 | 0.05 | 0.26 | 0.16 | **0.06** | **3.17E-04** | **0.14** |
| SM (d18:1/20:1, d18:2/20:0) | 0.08 | 0.56 | 0.09 | -2.97E-03 | 0.35 | 0.13 | 0.06 | 0.16 | 0.16 | 0.06 | 3.50E-04 | 0.14 |
| SM (d18:1/22:1, d18:2/22:0,<br>d16:1/24:1) | 0.15 | 0.19 | 0.10 | -3.62E-03 | 0.13 | 0.14 | 0.05 | 0.18 | 0.17 | 0.03 | 0.01 | 0.13 |
| SM (d18:1/24:1, d18:2/24:0) | 0.05 | 0.69 | 0.09 | -6.82E-04 | 0.79 | 0.14 | 0.01 | 0.73 | 0.16 | 0.03 | 0.04 | 0.13 |
| SM (d18:2/16:0, d18:1/16:1) | -0.08 | 0.57 | 0.10 | -4.76E-03 | 0.17 | 0.14 | 0.09 | 0.07 | 0.17 | **0.09** | **1.93E-06** | **0.14** |
| SM (d18:2/24:1, d18:1/24:2) | 0.06 | 0.59 | 0.09 | -4.02E-03 | 0.13 | 0.14 | 0.05 | 0.15 | 0.16 | 0.04 | 0.01 | 0.13 |
| stearoyl SM (d18:1/18:0) | -0.22 | 0.18 | 0.10 | -1.04E-03 | 0.84 | 0.14 | 0.05 | 0.50 | 0.16 | **0.37** | **1.83E-23** | **0.23** |

SM: sphingomyelin; PiB Global DVR: positron emission tomography Pittsburgh compound B; A $\beta_{42/40}$ : amyloid-beta 42/40 ratio; p-tau<sub>181</sub>/A $\beta_{42}$ : phosphorylated-tau<sub>181</sub> to amyloid-beta 42 ratio; p-tau<sub>181</sub>: phosphorylated-tau; $\beta$ : change in the outcome with a one unit increase of the standardized metabolite level; P: uncorrected P-value of the corresponding regression; R<sup>2</sup>: adjusted R<sup>2</sup> for PiB Global DVR, marginal R<sup>2</sup> for CSF biomarkers; bolded values indicate significant results at threshold $p < 3.47 \times 10^{-4}$ .

**Table 3.** Associations between SMs and neurodegeneration biomarkers.

| Metabolite | NfL (pg/mL) | | | α-synuclein (pg/mL) | | | Neurogranin (pg/mL) | | |
| --- | --- | --- | --- | --- | --- | --- | --- | --- | --- |
| | $\beta$ | P | R<sup>2</sup> | $\beta$ | P | R<sup>2</sup> | $\beta$ | P | R<sup>2</sup> |
| behenoyl SM (d18:1/22:0) | 0.06 | 1.23E-03 | 0.46 | 31.79 | 8.96E-04 | 0.09 | 46.86 | 0.08 | 0.04 |
| palmitoyl dihydro-SM (d18:0/16:0) | **0.15** | **2.14E-06** | **0.46** | **87.32** | **1.16E-07** | **0.11** | 148.66 | 0.01 | 0.05 |
| palmitoyl SM (d18:1/16:0) | **0.37** | **1.57E-18** | **0.52** | **254.75** | **1.72E-35** | **0.26** | **608.66** | **1.34E-15** | **0.10** |
| SM (d18:1/14:0, d16:1/16:0) | **0.16** | **8.83E-08** | **0.47** | **142.70** | **4.19E-18** | **0.16** | **336.80** | **1.74E-09** | **0.07** |
| SM (d18:1/18:1, d18:2/18:0) | **0.37** | **3.99E-17** | **0.51** | **247.38** | **3.30E-32** | **0.25** | **626.89** | **2.13E-15** | **0.10** |
| SM (d18:1/20:0, d16:1/22:0) | **0.17** | **2.34E-09** | **0.48** | **103.61** | **1.02E-12** | **0.12** | **207.71** | **8.00E-06** | **0.05** |
| SM (d18:1/20:1, d18:2/20:0) | **0.12** | **6.43E-05** | **0.45** | **64.18** | **3.29E-05** | **0.10** | 179.55 | 7.21E-04 | 0.05 |
| SM (d18:1/22:1, d18:2/22:0, d16:1/24:1) | **0.10** | **5.20E-06** | **0.46** | **50.48** | **1.34E-05** | **0.09** | 110.05 | 3.23E-03 | 0.05 |
| SM (d18:1/24:1, d18:2/24:0) | **0.09** | **1.30E-04** | **0.47** | **49.82** | **3.79E-05** | **0.09** | 81.38 | 0.04 | 0.04 |
| SM (d18:2/16:0, d18:1/16:1) | **0.22** | **5.16E-12** | **0.49** | **146.47** | **1.36E-19** | **0.17** | **337.93** | **1.39E-09** | **0.07** |
| SM (d18:2/24:1, d18:1/24:2) | **0.14** | **1.02E-08** | **0.48** | **75.29** | **2.36E-09** | **0.10** | 111.55 | 0.01 | 0.04 |
| stearoyl SM (d18:1/18:0) | **0.45** | **1.94E-21** | **0.53** | **325.23** | **1.25E-51** | **0.36** | **994.55** | **2.82E-28** | **0.18** |

SM: sphingomyelin; NfL: neurofilament light; $\beta$ : change in the outcome with a one unit increase of the standardized metabolite level; P: P-value of the corresponding regression;
R<sup>2</sup>: marginal R<sup>2</sup> for the corresponding regression; bolded values indicate significant results at threshold $p < 3.47 \times 10^{-4}$ .

**Figure 2.**
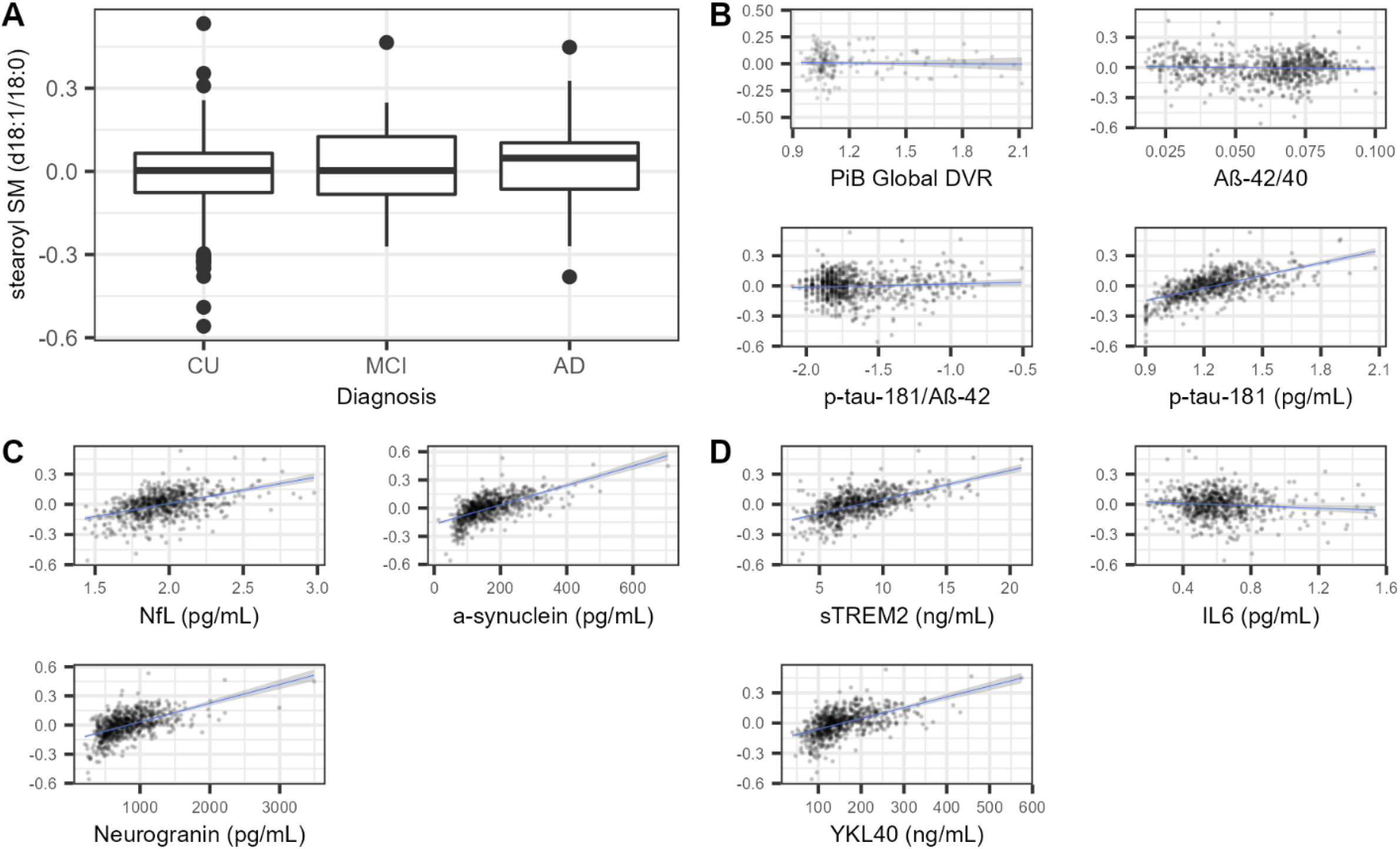
Associations between biomarkers, diagnosis, and stearoyl SM (d18:1/18:0). Results from stearoyl SM (d18:1/18:0) are displayed because this SM was consistently the most significantly associated SM and all other significant associations with SMs were in the same direction. **A)** Boxplots showing stearoyl SM (d18:1/18:0) levels for individuals with CU, MCI, and dementia-AD diagnoses. A small but statistically insignificant increase in metabolite level can be seen moving from the CU to AD group. None of the SM-diagnosis regressions were statistically significant. **B)** Scatterplots with each of the four AD-specific outcomes (x-axis) and stearoyl SM (d18:1/18:0) (y-axis). Stearoyl SM was not significantly associated with the measures of amyloid, but was significantly, positively associated with p-tau_181_ after Bonferroni correction (Table 2). **C)** Scatterplots of each of the three neurodegeneration biomarkers (x-axis) plotted against stearoyl SM (d18:1/18:0) (y-axis). Stearoyl SM was significantly, positively associated with NfL, neurogranin, and α-synuclein after Bonferroni correction (Table 3). **D)** Scatterplots of each of the three neuroinflammation biomarkers (x-axis) plotted against stearoyl SM (d18:1/18:0) (y-axis). Stearoyl SM was significantly, positively associated with YKL40 and sTREM2 and nominally, negatively associated with IL6 after Bonferroni correction (Table 4). **A-D:** The units of stearoyl SM (d18:1/18:0) as well as p-tau_181_, p-tau_181_/Aβ_42_, NfL, and IL6 are standardized by log_10_-transformation as laid out in the methods section of this paper. **B-D:** Longitudinal data were used to perform the regressions shown. Best fit lines constructed using linear regression models of form outcome ∼ stearoyl SM with 95% confidence intervals are drawn onto each of the scatterplots.

CSF levels of SMs were clearly associated with all CSF biomarkers of neurodegeneration. For all three biomarkers (neurogranin, NfL, α-synuclein), at least 11 of the 12 SMs were nominally and positively associated with each biomarker (Table 3). This positive association was visible in scatterplots of each outcome plotted against stearoyl SM, where there was a clear association and a consistent, positive trend (Figure 2C).

Associations between the 12 SMs and neuroinflammation biomarkers (YKL40, sTREM2, IL6) showed slightly less consistent results than the associations with the neurodegeneration biomarkers. Of the SM-IL6 associations, only three were nominally significant, and none were significant after multiple testing correction (Table 4 and Figure 2D). In contrast, 10 of the 12 SMs were nominally, positively associated with YKL40; 5 of these associations remained significant after Bonferroni correction. Finally, all 12 SMs had positive, nominally significant associations with sTREM2; 9 associations remained significant after Bonferroni correction.

**Table 4.** Associations between SMs and neuroinflammation biomarkers.

| Metabolite | sTREM2 (ng/mL) |  |  | IL6 (pg/mL) |  |  | YKL40 (ng/mL) |  |  |
| --- | --- | --- | --- | --- | --- | --- | --- | --- | --- |
| | $\beta$ | P | R<sup>2</sup> | $\beta$ | P | R<sup>2</sup> | $\beta$ | P | R<sup>2</sup> |
| behenoyl SM (d18:1/22:0) | 0.47 | 0.03 | 0.11 | -0.02 | 0.48 | 0.03 | 6.87 | 0.05 | 0.31 |
| palmitoyl dihydro-SM (d18:0/16:0) | **1.64** | **5.15E-05** | **0.12** | 0.02 | 0.75 | 0.03 | 19.80 | 3.67E-03 | 0.32 |
| palmitoyl SM (d18:1/16:0) | **4.55** | **1.06E-16** | **0.17** | -0.10 | 0.12 | 0.03 | **63.27** | **2.92E-11** | **0.35** |
| SM (d18:1/14:0, d16:1/16:0) | **1.91** | **6.67E-07** | **0.12** | -0.07 | 0.18 | 0.03 | **27.02** | **5.14E-05** | **0.33** |
| SM (d18:1/18:1, d18:2/18:0) | **4.30** | **2.70E-14** | **0.17** | -0.17 | 7.56E-03 | 0.04 | **59.56** | **2.40E-09** | **0.35** |
| SM (d18:1/20:0, d16:1/22:0) | **1.48** | **7.69E-06** | **0.12** | -0.06 | 0.22 | 0.03 | 15.09 | 0.01 | 0.32 |
| SM (d18:1/20:1, d18:2/20:0) | 0.88 | 0.01 | 0.12 | -0.01 | 0.78 | 0.02 | 13.51 | 0.03 | 0.33 |
| SM (d18:1/22:1, d18:2/22:0, d16:1/24:1) | **1.03** | **9.70E-05** | **0.11** | -0.03 | 0.40 | 0.03 | 9.82 | 0.03 | 0.32 |
| SM (d18:1/24:1, d18:2/24:0) | 0.92 | 7.96E-04 | 0.12 | -0.03 | 0.44 | 0.03 | 5.99 | 0.19 | 0.32 |
| SM (d18:2/16:0, d18:1/16:1) | **2.26** | **1.12E-08** | **0.13** | -0.14 | 8.31E-03 | 0.04 | **31.62** | **3.08E-06** | **0.33** |
| SM (d18:2/24:1, d18:1/24:2) | **1.28** | **1.27E-05** | **0.12** | -0.05 | 0.23 | 0.03 | 11.71 | 0.02 | 0.32 |
| stearoyl SM (d18:1/18:0) | **6.99** | **3.15E-28** | **0.24** | -0.18 | 5.77E-03 | 0.04 | **105.64** | **1.41E-18** | **0.39** |

*sTREM2: soluble triggering receptor found on myeloid cells 2; IL6: interleukin-6; YKL40: chitinase-3-like protein 1; $\beta$ : change in the outcome with a one unit increase of the standardized metabolite level; P: P-value of the corresponding regression; R<sup>2</sup>: marginal R<sup>2</sup> for the corresponding regression; bolded values indicate significant results at threshold $p < 3.47 \times 10^{-4}$ .*

Behenoyl SM (d18:1/22:0) yielded no significant associations after Bonferroni correction with any biomarkers analyzed, while stearoyl SM (d18:1/18:0) had the strongest significant association with all biomarkers where any significant association was observed. Palmitoyl SM (d18:1/16:0), SM (d18:1/14:0, d16:1/16:0), SM (d18:1/18:1, d18:2/18:0), SM (d18:2/16:0, d18:1/16:1), and stearoyl SM (d18:1/18:0) were significantly associated with the same seven biomarkers (p-tau_181_, NfL, α**-**synuclein, neurogranin, sTREM2, IL6, and YKL40) after Bonferroni correction.

### 3.4. Independent signals

Six models with p-tau_181_ as the outcome and stearoyl SM (the most significantly associated metabolite) as the main predictor were significantly improved by the addition of another metabolite predictor using nested ANOVA models (Supplemental Table 6). Only one metabolite significantly improved the model for NfL, while there were six metabolites that significantly improved the model for α-synuclein and seven that significantly improved the model for neurogranin. Among the models for neuroinflammation biomarkers, three metabolites significantly improved the models for sTREM2, one significantly improved the model for IL6, and five metabolites significantly improved the model for YKL40.

### 3.5. Sensitivity Analyses

The results of the sensitivity analysis with the *APOE* ε4 covariate were compared with the main analysis results and were not substantially different in either significance of associations or direction of effect (Supplemental Table 7). Sensitivity analyses testing for sex-specific differences in associations between SMs and the outcomes found no significant interaction effects after Bonferroni correction either when testing for the significance of an interaction term in the regression (Supplemental Table 8). or when stratifying by sex (Supplemental Table 9). Sensitivity analyses testing for modification of the effect of SMs on the outcomes by amyloid and tau status suggested several significant differences. Simple main effects from the interaction testing suggested that effects of SMs were stronger in the A+T+ group for some biomarker and SM combinations, without a strong pattern. Five SM-neurodegeneration biomarker regressions and nine SM-neuroinflammation biomarker regressions had significant A+T+ interaction effects, in all cases having a positive interaction effect term indicating the biomarker level rising more quickly per unit increase in the SMs among the A+T+ group. Notably, 7 of the 12 SMs had a positive, significant interaction term in the models for YKL-40 (Supplemental Table 10 Supplemental Figure 2). Similarly, in the amyloid and tau (AT)-stratified regressions, there were generally larger effect sizes observed in the A+T+ group than the A-T-group for the biomarkers NfL, α-synuclein, neurogranin, sTREM2, and YKL40 when there were significant effects detected (Supplemental Table 11), though the smaller sample sizes among the subgroups likely lowered the power for detecting some of the SM-biomarker associations.

## 4. Discussion

Our study provides a robust exploration of the relationship between CSF SMs and various aspects of AD. We first explored whether CSF SMs were associated with diagnosis of AD or MCI in our cohorts. We found no significant associations between SMs and diagnosis of AD or MCI, partly contrasting with previous work that found an association between CSF SMs and prodromal AD, though not with mild or moderate AD[41]. The smaller sample size (n = 37) in the previous study may account for this difference, though it could also be that the differences in the particular subgroups of AD studied (CU/MCI/AD vs controls/prodromal/mild/moderate AD) led to this particular early stage signal going undetected in our analyses. Similarly, we found that amyloid biomarkers (Aβ_42/40_, PiB Global DVR, and p-tau_181_/Aβ_42_) were not significantly associated with the CSF SMs analyzed. This finding is consistent with previous research in the WRAP cohort that found no association between any of the CSF metabolites and these measures of amyloid[14]. However, there were significant SM associations with p-tau_181_ and several biomarkers for neurodegeneration and neuroinflammation. Five SMs (stearoyl SM (d18:1/18:0), SM (d18:2/16:0, d18:1/16:1), SM (d18:1/18:1, d18:2/18:0), SM (d18:1/14:0, d16:1/16:0), and palmitoyl SM (d18:1/16:0)) in particular were significantly, positively associated with p-tau_181_ and each of the biomarkers for neurodegeneration (NfL, α-synuclein, and neurogranin). The same five SMs were significantly, positively associated with the neuroinflammation markers YKL40 and sTREM2. All these associations were similar when we controlled for the number of *APOE* ε4 alleles, indicating that these results were not likely to be influenced by *APOE* genotype. Of the twelve metabolites we analyzed, stearoyl SM (d18:1/18:0) had the strongest associations with our biomarkers when there was any statistically significant SM association observed. This particular metabolite, along with SM (18:1/18:1), has been previously identified as having a significant, positive association with AD pathology[42]. While neither of these metabolites were significantly associated with markers of amyloid in our study, they were both associated with p-tau_181_ and multiple biomarkers of neurodegeneration and neuroinflammation. Our pairwise correlations indicate that many of the CSF SMs we analyzed were intercorrelated, with potential subgroups among SMs based on chain length. Nested linear models with our most strongly associated metabolite, stearoyl SM (d18:1/18:0), supported the correlated nature of the SMs. Many of these models also showed some improvement with the addition of certain SMs, indicating that SMs other than stearoyl SM (d18:1/18:0) contain additional useful predictive information.

In the secondary analyses, we found no significant effect modifications by sex and no difference between AT groups in relationships between SMs and cognitive status. However, we found evidence suggesting stronger SM effects on several biomarker outcomes, including α-synuclein, neurogranin, sTREM2, and YKL40, within the A+T+ subgroup than in other subgroups. Larger observed effect sizes in the A+T+ groups occurred for the biomarkers generally expected to change later on in the disease trajectory (e.g. p-tau)[43]. More research with larger sample sizes will be necessary to establish associations between SMs and various stages of AD more clearly given the relatively smaller sample sizes in the A+T- and A+T+ groups in this study.

Collectively, our results implicate CSF SMs as non-specific biomarkers of neurodegeneration and neuroinflammation. Strong positive associations between many SMs and markers like p-tau_181_, NfL, neurogranin, YKL-40, and sTREM2 make SMs a candidate marker of neurodegenerative and neuroinflammatory processes, but the lack of association of these SMs with amyloid measures might mean that SMs’ changes are not specific to AD. One potential mechanism for the role of SMs in neurodegeneration and neuroinflammation is that SMs only begin changing as neurodegeneration or neuroinflammation occur and neurons begin to die, but not earlier, when amyloid is first beginning to accumulate. Previous research in preeclampsia has highlighted a mechanism by which SM levels increase. Stearoyl SM (d18:1/18:0), along with palmitoyl SM (d18:1/16:0), was elevated in plasma of preeclamptic mothers, thought to be a result of lipid rafts’ (microdomains of the cell membrane, consisting of SMs and other compounds) exposure to low oxygen levels[44]. This finding may grant insight into potential mechanisms for the association of CSF SMs with neurodegeneration: as neurons die, lipid rafts might release their components, including SMs, into CSF[8,10]. Mechanistic research into the SM changes during neurodegeneration are needed to further examine these hypotheses.

As noted above, previous studies on the relationship between AD and SMs in plasma and brain tissue have found significant associations but differing directionality, making CSF SMs especially interesting [9,10]. Our lack of significant SM-amyloid associations contrasted with a previous study on the association of total SMs with Aβ in CSF, which found significant associations between SMs (including stearoyl SM (d18:0/18:1)) and various markers of amyloid including Aβ_42_ and Aβ_40_[45]. Here, with a larger sample size, we found no association of SMs with Aβ markers. A few differences could explain this discrepancy, including differences in study population (e.g., our population was enriched for older individuals, mean = 64.1) or different biomarkers used (e.g., Aβ_42_ and Aβ_40_ vs Aβ_42_/Aβ_40_).

This study has certain limitations that must be taken into consideration when interpreting its results. While we were able to produce one of the largest sample sizes of AD and MCI individuals used to study CSF SMs, our samples of individuals with AD and MCI were still relatively small (n = 89) compared to cognitively unimpaired controls (n = 409) (Table 1), potentially limiting our ability to assess diagnosis-related outcomes. We also lacked diversity; over 95% of individuals included in this study were self-reported white, limiting the generalizability of this study to other populations. As sample sizes for underrepresented groups grow, we will be better able to investigate the role of SMs across a greater range of populations. We also were limited by the types of sphingolipids that we examined.

Ceramides, particularly, are a type of sphingolipid that has been implicated in AD and other diseases, but we were not able to assess their role here, as they were not measured in CSF for our study[46]. There is a high correlation between p-tau_181_ and neurodegeneration biomarkers in this cohort, which may differ in a sample with more non-AD dementia present. Finally, nine SMs analyzed were designated as tier 2 compounds by Metabolon, compounds that have a structure that has been confirmed by literature review, but they are not necessarily confirmed by a reference standard, though we note that stearoyl SM (d18:1/18:0) was not one of these tier 2 compounds[47].

## 5. Conclusion

In this study, we examined the relationship between 12 SMs, MCI/AD diagnoses, and PET and CSF biomarkers of AD, neurodegeneration, and neuroinflammation, providing a comprehensive investigation of the role of CSF SMs in AD pathology. While we found no association between SMs and dementia-AD or MCI diagnoses nor with amyloid biomarkers, we did find strong positive associations between the SMs and p-tau_181_, NfL, sTREM2, neurogranin, α-synuclein, and YKL40. Based on these findings, we hypothesize that SMs are non-specific biomarkers of neurodegeneration and neuroinflammation. There is still much to be learned: identifying a mechanism for SMs’ role in neurodegeneration and neuroinflammation, understanding how SM levels differ across other neurodegenerative diseases, and characterizing them in diverse cohorts with larger numbers of AD individuals will grant greater insights into the role of these metabolites in AD.

## Data Availability

The datasets analyzed in this study may be requested from the Wisconsin ADRC at https://www.adrc.wisc.edu/apply-resources.

## Abbreviations

Aβ: amyloid beta
AD: Alzheimer’s disease
Dementia-AD: clinical diagnosis of dementia with AD as a suspected cause
ADRC: Alzheimer’s Disease Research Center
α-synuclein: alpha synuclein
ANOVA: analysis of variance
CSF: cerebrospinal fluid
CU: cognitively unimpaired
DVR: distribution volume ratio
IL6: interleukin-6
LMM: linear mixed effects model
LP: lumbar puncture
MCI: mild cognitive impairment
MRI: magnetic resonance imaging
NfL: neurofilament light
NTK: NeuroToolKit
PET: positron emission tomography
PiB: Pittsburgh compound B
p-tau_181_: phosphorylated tau
p-tau_181_/Aβ_42_: p-tau_181_ to amyloid beta 42 ratio
QC: quality control
ROI: region of interest
SM: sphingomyelin
sTREM2: soluble triggering receptor found on myeloid cells 2
WRAP: Wisconsin Registry for Alzheimer’s Prevention
YKL40: chitinase-3-like protein 1

## Acknowledgements

We thank the participants in the WRAP and WADRC studies and the staff of the Wisconsin Alzheimer’s Institute and WADRC; it is because of these individuals that this research is possible. We also would like to thank all members of the Engelman lab for their feedback and support of this research.

## Funding Sources

This research is supported by National Institutes of health (NIH) grants R01AG27161 (WRAP: Biomarkers of Preclinical AD), R01AG054047 (Genomic and Metabolomic Data Integration in a Longitudinal Cohort at Risk for AD), P30AG017266 (Center for Demography of Health and Aging), P50AG033514 and P30AG062715 (Wisconsin ADRC Grant), R01 AG021155 (Longitudinal Course of Imaging Biomarkers in People At Risk for AD); UL1TR000427 (Clinical and Translational Science Award (CTSA) program through the NIH National Center for Advancing Translational Sciences (NCATS), P2CHD047873 (core grant to the Center for Demography and Ecology at the University of Wisconsin-Madison, S10 OD025245-01 (Biomedical Research Support Shared Instrumentation grant from NIH). Author YKD was supported by a training grant from the National Institute on Aging (T32AG000213). Author HZ is a Wallenberg Scholar supported by grants from the Swedish Research Council (#2018-02532), the European Research Council (#681712), Swedish State Support for Clinical Research (#ALFGBG-720931), the Alzheimer Drug Discovery Foundation (ADDF), USA (#201809-2016862), the AD Strategic Fund and the Alzheimer’s Association (#ADSF-21-831376-C, #ADSF-21-831381-C and #ADSF-21-831377-C), the Olav Thon Foundation, the Erling-Persson Family Foundation, Stiftelsen för Gamla Tjänarinnor, Hjärnfonden, Sweden (#FO2019-0228), the European Union’s Horizon 2020 research and innovation programme under the Marie Skłodowska-Curie grant agreement No 860197 (MIRIADE), and the UK Dementia Research Institute at UCL. Author KB is supported by the Swedish Research Council (#2017-00915), the Alzheimer Drug Discovery Foundation (ADDF), USA (#RDAPB-201809-2016615), the Swedish Alzheimer Foundation (#AF-742881), Hjärnfonden, Sweden (#FO2017-0243), the Swedish state under the agreement between the Swedish government and the County Councils, the ALF-agreement (#ALFGBG-715986), the European Union Joint Program for Neurodegenerative Disorders (JPND2019-466-236), the National Institute of Health (NIH), USA, (grant #1R01AG068398-01), and the Alzheimer’s Association 2021 Zenith Award (ZEN-21-848495). Author TJB and PET data processing and analyses are supported by the Alzheimer’s Association (AARF-19-614533). Author EV was supported by the Center for Demography of Health and Aging (P30 AG17266) and a NIA Training Grant (Population, Life Course and Aging) (T32 AG00129).

The Roche NeuroToolKit is a panel of robust prototype biomarker assays designed to evaluate key pathologic events characteristic of AD and other neurological disorders, used for research purposes only and not approved for clinical use.

## Notes

### Competing Interest Statement

Author EJ's spouse is an employee of, and owns stock options in, Epic. HZ, author, has served at scientific advisory boards and/or as a consultant for Alector, Eisai, Denali, Roche Diagnostics, Wave, Samumed, Siemens Healthineers, Pinteon Therapeutics, Nervgen, AZTherapies, CogRx and Red Abbey Labs, has given lectures in symposia sponsored by Cellectricon, Fujirebio, Alzecure and Biogen, and is a co-founder of Brain Biomarker Solutions in Gothenburg AB (BBS), which is a part of the GU Ventures Incubator Program (outside submitted work). KB has served as a consultant, at advisory boards, or at data monitoring committees for Abcam, Axon, Biogen, JOMDD/Shimadzu. Julius Clinical, Lilly, MagQu, Novartis, Prothena, Roche Diagnostics, and Siemens Healthineers, and is a co-founder of Brain Biomarker Solutions in Gothenburg AB (BBS), which is a part of the GU Ventures Incubator Program, all unrelated to the work presented in this paper. SCJ, author, has served at advisory boards for Roche Diagnostics and was a consultant to Roche Diagnostics in 2018. Authors GK and AB are employees of Roche Diagnostics GmbH; AB owns stock in Hoffmann-La Roche Ltd. Author IS is an employee of Roche Diagnostics International Ltd.

### Author Declarations

This study was approved by the University of Wisconsin Health Sciences Institutional Review Board as part of the Generations of WRAP (GROW) study. Participants in the WADRC and WRAP studies provided written informed consent.

## References

[1] Wishart DS. Emerging applications of metabolomics in drug discovery and precision medicine. Nat Rev Drug Discov 2016;15:473–84. https://doi.org/10.1038/nrd.2016.32.

[2] Bain JR, Stevens RD, Wenner BR, Ilkayeva O, Muoio DM, Newgard CB. Metabolomics applied to diabetes research: moving from information to knowledge. Diabetes 2009;58:2429–43. https://doi.org/10.2337/db09-0580.

[3] Puchades-Carrasco L and Pineda-Lucena A. Metabolomics Applications in Precision Medicine: An Oncological Perspective. Curr Top Med Chem 2017;17:2740–51. https://doi.org/10.2174/1568026617666170707120034.

[4] Metabolomic analysis of the cerebrospinal fluid reveals changes in phospholipase expression in the CNS of SIV-infected macaques. - Abstract - Europe PMC n.d. https://europepmc.org/article/pmc/pmc2398736 (accessed August 3, 2020).

[5] The Role of Metabolomics in Brain Metabolism Research. - Abstract - Europe PMC n.d. https://europepmc.org/article/med/26201839 (accessed August 3, 2020).

[6] Wilkins JM, Trushina E. Application of Metabolomics in Alzheimer’s Disease. Front Neurol 2018;8. https://doi.org/10.3389/fneur.2017.00719.

[7] Han X, Rozen S, Boyle SH, Hellegers C, Cheng H, Burke JR, et al. Metabolomics in early Alzheimer’s disease: identification of altered plasma sphingolipidome using shotgun lipidomics. PloS One 2011;6:e21643. https://doi.org/10.1371/journal.pone.0021643.

[8] Pralhada Rao R, Vaidyanathan N, Rengasamy M, Mammen Oommen A, Somaiya N, Jagannath MR. Sphingolipid Metabolic Pathway: An Overview of Major Roles Played in Human Diseases. J Lipids 2013. https://doi.org/10.1155/2013/178910.

[9] Mielke MM, Lyketsos CG. Alterations of the sphingolipid pathway in Alzheimer’s disease: new biomarkers and treatment targets? Neuromolecular Med 2010;12:331–40. https://doi.org/10.1007/s12017-010-8121-y.

[10] Crivelli SM, Giovagnoni C, Visseren L, Scheithauer A-L, de Wit N, den Hoedt S, et al. Sphingolipids in Alzheimer’s disease, how can we target them? Adv Drug Deliv Rev 2020. https://doi.org/10.1016/j.addr.2019.12.003.

[11] Johnson SC, Koscik RL, Jonaitis EM, Clark LR, Mueller KD, Berman SE, et al. The Wisconsin Registry for Alzheimer’s Prevention: A review of findings and current directions. Alzheimers Dement Diagn Assess Dis Monit 2017;10:130–42. https://doi.org/10.1016/j.dadm.2017.11.007.

[12] Melah KE, Lu SY-F, Hoscheidt SM, Alexander AL, Adluru N, Destiche DJ, et al. CSF markers of Alzheimer’s pathology and microglial activation are associated with altered white matter microstructure in asymptomatic adults at risk for Alzheimer’s disease. J Alzheimers Dis JAD 2016;50:873–86. https://doi.org/10.3233/JAD-150897.

[13] Hulle CV, Jonaitis EM, Betthauser TJ, Batrla R, Wild N, Kollmorgen G, et al. An examination of a novel multipanel of CSF biomarkers in the Alzheimer’s disease clinical and pathological continuum. Alzheimers Dement 2021;17:431–45. https://doi.org/10.1002/alz.12204.

[14] Darst BF, Lu Q, Johnson SC, Engelman CD. Integrated analysis of genomics, longitudinal metabolomics, and Alzheimer’s risk factors among 1,111 cohort participants. Genet Epidemiol 2019;43:657–74. https://doi.org/10.1002/gepi.22211.

[15] Johnson SC, Christian BT, Okonkwo OC, Oh JM, Harding S, Xu G, et al. Amyloid burden and neural function in people at risk for Alzheimer’s Disease. Neurobiol Aging 2014;35:576–84. https://doi.org/10.1016/j.neurobiolaging.2013.09.028.

[16] SPM - Statistical Parametric Mapping n.d. https://www.fil.ion.ucl.ac.uk/spm/ (accessed September 3, 2021).

[17] Tzourio-Mazoyer N, Landeau B, Papathanassiou D, Crivello F, Etard O, Delcroix N, et al. Automated anatomical labeling of activations in SPM using a macroscopic anatomical parcellation of the MNI MRI single-subject brain. NeuroImage 2002;15:273–89. https://doi.org/10.1006/nimg.2001.0978.

[18] Lopresti BJ, Klunk WE, Mathis CA, Hoge JA, Ziolko SK, Lu X, et al. Simplified Quantification of Pittsburgh Compound B Amyloid Imaging PET Studies: A Comparative Analysis. J Nucl Med 2005;46:1959–72.

[19] Janelidze S, Zetterberg H, Mattsson N, Palmqvist S, Vanderstichele H, Lindberg O, et al. CSF Aβ42/Aβ40 and Aβ42/Aβ38 ratios: better diagnostic markers of Alzheimer disease. Ann Clin Transl Neurol 2016;3:154– 65. https://doi.org/10.1002/acn3.274.

[20] Fagan AM, Roe CM, Xiong C, Mintun MA, Morris JC, Holtzman DM. Cerebrospinal fluid tau/beta-amyloid(42) ratio as a prediction of cognitive decline in nondemented older adults. Arch Neurol 2007;64:343– 9. https://doi.org/10.1001/archneur.64.3.noc60123.

[21] Buerger K, Ewers M, Pirttilä T, Zinkowski R, Alafuzoff I, Teipel SJ, et al. CSF phosphorylated tau protein correlates with neocortical neurofibrillary pathology in Alzheimer’s disease. Brain J Neurol 2006;129:3035–41. https://doi.org/10.1093/brain/awl269.

[22] Portelius E, Zetterberg H, Skillbäck T, Törnqvist U, Andreasson U, Trojanowski JQ, et al. Cerebrospinal fluid neurogranin: relation to cognition and neurodegeneration in Alzheimer’s disease. Brain 2015;138:3373–85. https://doi.org/10.1093/brain/awv267.

[23] Dhiman K, Gupta VB, Villemagne VL, Eratne D, Graham PL, Fowler C, et al. Cerebrospinal fluid neurofilament light concentration predicts brain atrophy and cognition in Alzheimer’s disease. Alzheimers Dement Diagn Assess Dis Monit 2020;12:e12005. https://doi.org/10.1002/dad2.12005.

[24] Majbour NK, Chiasserini D, Vaikath NN, Eusebi P, Tokuda T, van de Berg W, et al. Increased levels of CSF total but not oligomeric or phosphorylated forms of alpha-synuclein in patients diagnosed with probable Alzheimer’s disease. Sci Rep 2017;7:40263. https://doi.org/10.1038/srep40263.

[25] Kim YS, Lee KJ, Kim H. Serum tumour necrosis factor-α and interleukin-6 levels in Alzheimer’s disease and mild cognitive impairment. Psychogeriatr Off J Jpn Psychogeriatr Soc 2017;17:224–30. https://doi.org/10.1111/psyg.12218.

[26] Nordengen K, Kirsebom B-E, Henjum K, Selnes P, Gísladóttir B, Wettergreen M, et al. Glial activation and inflammation along the Alzheimer’s disease continuum. J Neuroinflammation 2019;16:46. https://doi.org/10.1186/s12974-019-1399-2.

[27] Suárez-Calvet M, Morenas-Rodríguez E, Kleinberger G, Schlepckow K, Araque Caballero MÁ, Franzmeier N, et al. Early increase of CSF sTREM2 in Alzheimer’s disease is associated with tau related-neurodegeneration but not with amyloid-β pathology. Mol Neurodegener 2019;14:1. https://doi.org/10.1186/s13024-018-0301-5.

[28] Joanes DN, Gill CA. Comparing measures of sample skewness and kurtosis. J R Stat Soc Ser Stat 1998;47:183–9. https://doi.org/10.1111/1467-9884.00122.

[29] Tidyverse n.d. https://www.tidyverse.org/ (accessed August 31, 2020).

[30] stats package | R Documentation n.d. https://www.rdocumentation.org/packages/stats/versions/3.6.2 (accessed September 20, 2020).

[31] Bates D, Mächler M, Bolker B, Walker S. Fitting Linear Mixed-Effects Models Using lme4. J Stat Softw 2015;67:1–48. https://doi.org/10.18637/jss.v067.i01.

[32] Kuznetsova A, Brockhoff PB, Christensen RHB. lmerTest Package: Tests in Linear Mixed Effects Models. ÁJ Stat Softw 2017;82:1–26. https://doi.org/10.18637/jss.v082.i13.

[33] Murphy M. semEff: Automatic Calculation of Effects for Piecewise Structural Equation Models. 2020.

[34] r.squaredGLMM function | R Documentation n.d. https://www.rdocumentation.org/packages/MuMIn/versions/1.40.4/topics/r.squaredGLMM (accessed October 31, 2020).

[35] Liu C-C, Kanekiyo T, Xu H, Bu G. Apolipoprotein E and Alzheimer disease: risk, mechanisms, and therapy. Nat Rev Neurol 2013;9:106–18. https://doi.org/10.1038/nrneurol.2012.263.

[36] Kulminski AM, Shu L, Loika Y, Nazarian A, Arbeev K, Ukraintseva S, et al. APOE region molecular signatures of Alzheimer’s disease across races/ethnicities. Neurobiol Aging 2020;87:141.e1-141.e8. https://doi.org/10.1016/j.neurobiolaging.2019.11.007.

[37] Mielke MM, Haughey NJ, Han D, An Y, Bandaru VVR, Lyketsos CG, et al. The Association Between Plasma Ceramides and Sphingomyelins and Risk of Alzheimer’s Disease Differs by Sex and APOE in the Baltimore Longitudinal Study of Aging. J Alzheimers Dis JAD 2017;60:819–28. https://doi.org/10.3233/JAD-160925.

[38] Pujol-Lereis LM. Alteration of Sphingolipids in Biofluids: Implications for Neurodegenerative Diseases. Int J Mol Sci 2019;20:3564. https://doi.org/10.3390/ijms20143564.

[39] Darst BF, Koscik RL, Hogan KJ, Johnson SC, Engelman CD. Longitudinal plasma metabolomics of aging and sex. Aging 2019;11:1262–82. https://doi.org/10.18632/aging.101837.

[40] NIA□AA Research Framework: Toward a biological definition of Alzheimer’s disease - Jack - 2018 - Alzheimer’s &amp; Dementia - Wiley Online Library n.d. https://alz-journals.onlinelibrary.wiley.com/doi/full/10.1016/j.jalz.2018.02.018 (accessed September 14, 2020).

[41] Kosicek M, Zetterberg H, Andreasen N, Peter-Katalinic J, Hecimovic S. Elevated cerebrospinal fluid sphingomyelin levels in prodromal Alzheimer’s disease. Neurosci Lett 2012;516:302–5. https://doi.org/10.1016/j.neulet.2012.04.019.

[42] Koal T, Klavins K, Seppi D, Kemmler G, Humpel C. Sphingomyelin SM(d18:1/18:0) is significantly enhanced in cerebrospinal fluid samples dichotomized by pathological amyloid-β42, tau, and phospho-tau-181 levels. J Alzheimers Dis JAD 2015;44:1193–201. https://doi.org/10.3233/JAD-142319.

[43] Jack CR, Knopman DS, Jagust WJ, Petersen RC, Weiner MW, Aisen PS, et al. Update on hypothetical model of Alzheimer’s disease biomarkers. Lancet Neurol 2013;12:207–16. https://doi.org/10.1016/S1474-4422(12)70291-0.

[44] Ermini L, Ausman J, Melland-Smith M, Yeganeh B, Rolfo A, Litvack ML, et al. A Single Sphingomyelin Species Promotes Exosomal Release of Endoglin into the Maternal Circulation in Preeclampsia. Sci Rep 2017;7:12172. https://doi.org/10.1038/s41598-017-12491-4.

[45] Mielke MM, Haughey NJ, Bandaru VVR, Zetterberg H, Blennow K, Andreasson U, et al. CSF sphingolipids, β-amyloid, and tau in adults at risk for Alzheimer’s disease. Neurobiol Aging 2014;35:2486–94. https://doi.org/10.1016/j.neurobiolaging.2014.05.019.

[46] Czubowicz K, Jesko H, Wencel P, Lukiw WJ, Strosznajder RP. The Role of Ceramide and Sphingosine-1-Phosphate in Alzheimer’s Disease and Other Neurodegenerative Disorders. Mol Neurobiol 2019;56:5436–55. https://doi.org/10.1007/s12035-018-1448-3.

[47] Tier 1 Metabolite Identifications: A Compass to Rich Research Insights. Metabolon 2019. https://metabolon.com/tier-1-metabolite-identifications-a-compass-to-rich-research-insights/ (accessed December 22, 2020).

