## Supplemental Figures for "CSF sphingomyelins in Alzheimer’s disease, neurodegeneration, and neuroinflammation"

**Supplemental Figure 1**: A diagram of the sphingolipid metabolic pathway.

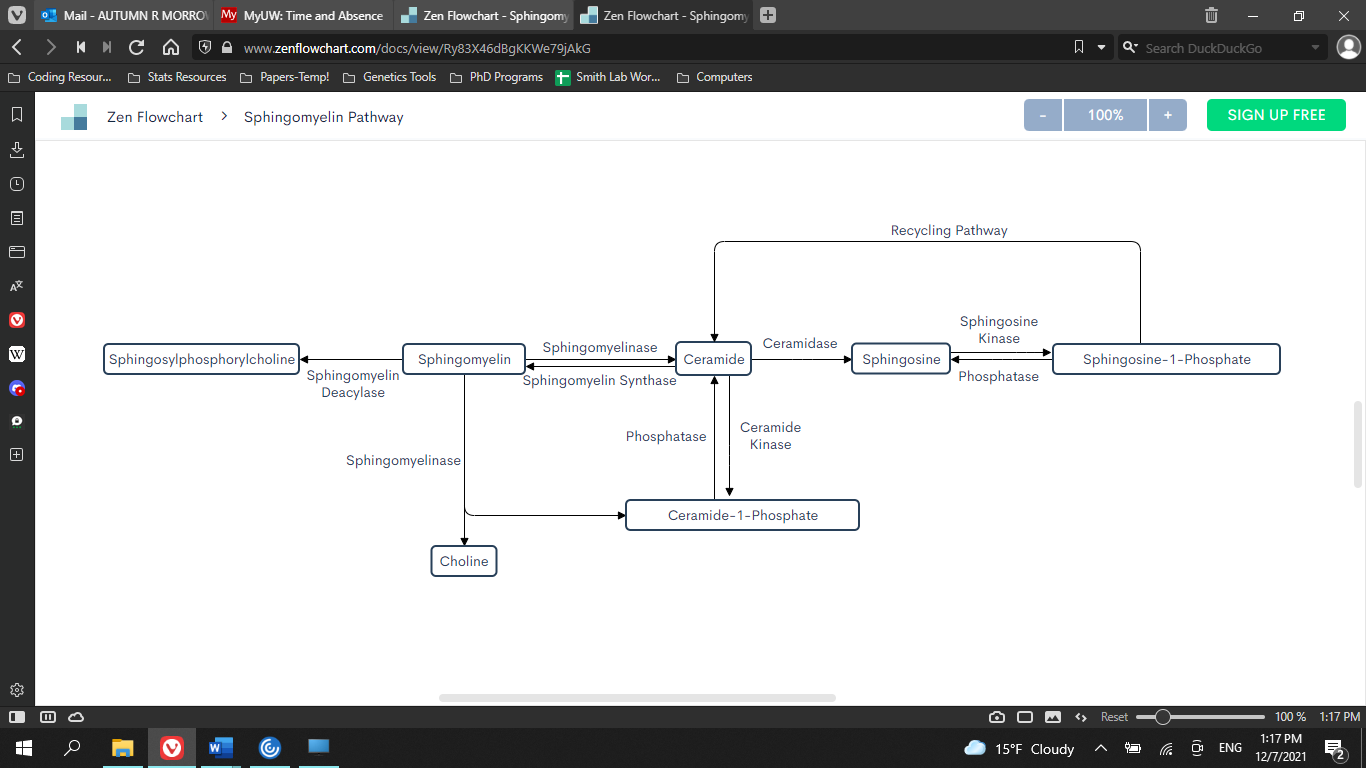

Sphingomyelins can be directly synthesized from or into ceramides and are involved in the synthesis of sphingosylphosphorylcholine in addition to ceramide-1-phosphate.

**Supplemental Figure 2:** Plots of stearoyl SM’s interaction effects by AT status.

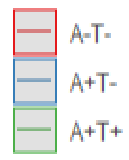

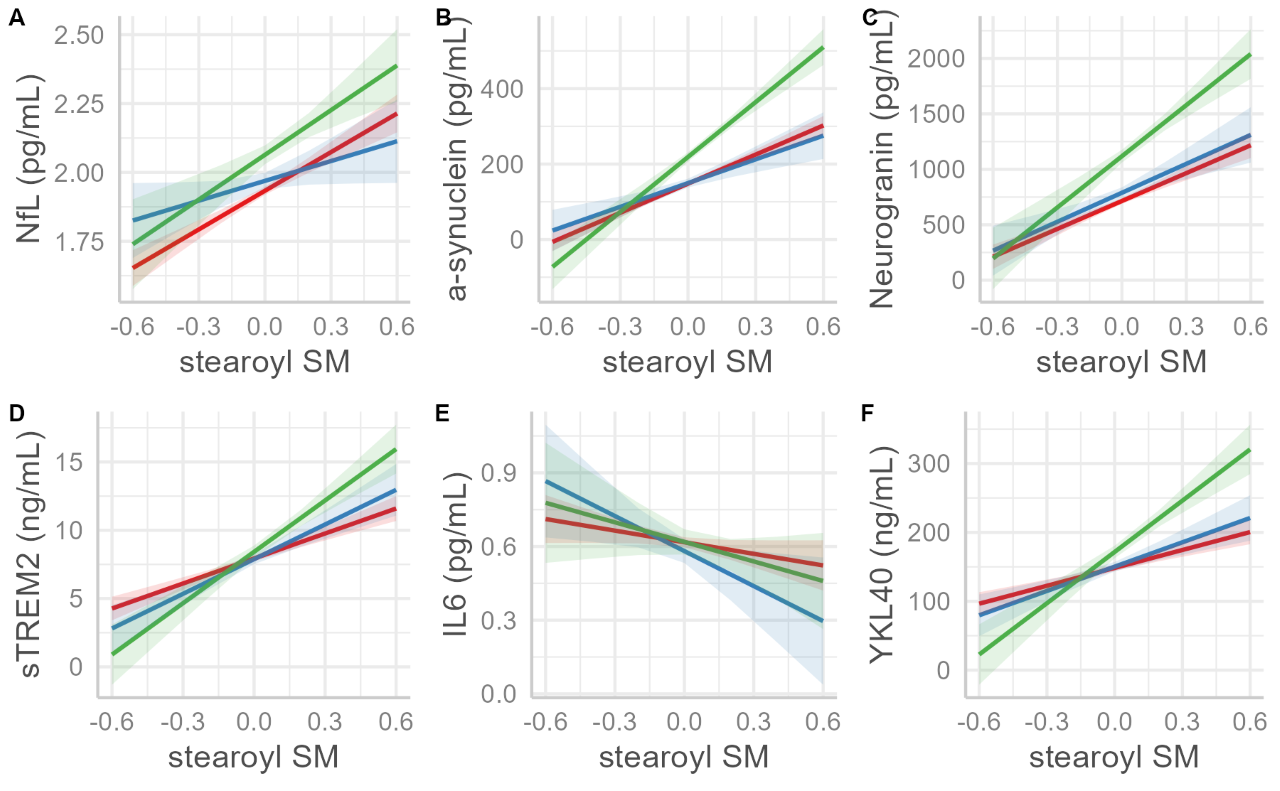

The interaction effects (with confidence intervals) of AT status are displayed for each outcome paired with stearoyl SM. Significant interactions (P < 3.47 × 10^-4^) with A+T+ status were observed for models in which α-synuclein, YKL40, and sTREM2 were regressed on stearoyl SM (x-axis).
